# Neuroticism levels in severe depression may reveal association with higher levels of intelligence

**DOI:** 10.1101/2023.11.22.23298698

**Authors:** Frederick Nitchie, Tyler Moore, J. Cobb Scott, Walid Makhoul, Yvette Sheline

## Abstract

The anxious-misery spectrum of disorders encompasses some of the most debilitating mental disorders that effect the population at-large. In addition, neuroticism is a trait that has been shown to correlate with higher levels of reported symptomatology for individuals diagnosed with anxious-misery disorders. However, it is less understood which of these might be more predictive of the cognitive impairments typically associated with mental illness. Therefore, we selected a group of participants with high trait neuroticism and comorbid anxious-misery symptoms. Using confirmatory factor analysis, we loaded the results of seventeen neuropsychological measures from each participant onto five distinct factors that exists on the anxious-misery spectrum: Depression, Anxiety, Impulsivity, Insomnia, and Somatizing. These factors were then compared to the results that participants had on to neurocognitive batteries: the University of Pennsylvania Computerized Neurocognitive Test Battery as well and the National Institute of Health Toolbox. Contrary to our expectations, we found that those in the top quartile of depressive symptom severity performed better than controls on working memory measures. These results are thought to be the result of correlations that have been shown previously between high levels of neuroticism and higher levels of intelligence.

## Introduction

In the field of mental health research, the term “anxious misery” has gained widespread use when referring to anxious and depressive disorders (Kreuger, 1999; Watson et al., 2008). These disorders include major depressive disorder (MDD), generalized anxiety disorder (GAD), and post-traumatic stress disorder (PTSD), among others. Symptoms common to these disorders are ruminative thoughts, negative mood states, feelings of nervousness and/or fear, and general difficulty in starting or maintaining effortful activity. These anxious-misery disorders display a large degree of overlap, both in symptomatology as well as comorbidity in those diagnosed with at least one disorder. Symptoms also display a tendency to persist even when comorbid symptoms are assessed and treated, leading to the hypothesis that the categorical dimensions of traditional psychiatric diagnosis may fail to encompass the full scope and severity of the underlying dysfunction.

Due to these overlapping symptoms and frequent co-occurrence of diagnoses, it has become increasingly common to think of these disorders less as distinctive entities and more as component groupings of symptoms that fall under a larger, overarching construct known as the Negative Valence System (NVS). The NVS is composed of many factors, with two of the most central factors being “responses to sustained threat” and “loss” (Hasratian et al., 2022). ‘Responses to sustained threat’ encompasses factors such as increased conflict detection, anxious arousal, attentional bias to threat, and avoidance. ‘Loss’ comprises factors such as rumination, sadness, guilt, anhedonia, withdrawal, negatively valanced attentional bias, and deficits in executive functioning. These types of symptoms are well-known to highly correlate with increased levels of neuroticism within individuals (Subica et al., 2016).

Neuroticism is generally defined as a tendency for one to react with negative emotionality to various sources of stress (Barlow et al., 2014). The negative emotions typically expressed include anxiety, fear, anger, and sadness. Due to this, neuroticism is generally seen as a precursor or risk factor to developing a mental disorder, especially those on the anxious-misery spectrum. Notably, these symptoms overlap largely with the ‘loss’ and ‘response to sustained threat’ factors of the NVS. What is less clear however, is which of these metrics (neuroticism scores vs clinical diagnoses) is more predictive of performance on typical measures of cognition.

We collected a large dataset composed of individuals who were selected for high levels of neuroticism on the Neuroticism-Extraversion-Openness Five-Factor Inventory (NEOFFIN). All participants were also diagnosed via structured clinical interview (SCID-V) to determine if they met criteria for the diagnoses in question (MDD, GAD, PTSD). We were interested in seeing how these levels of neuroticism and symptomatology correlated with performance on multiple cognitive domains and to determine whether neuroticism or clinical diagnosis was more predictive of performance.

## Methods

### Participants

#### Inclusion/Exclusion criteria

Participants were included if they: (i) were between 18 and 59 years of age; (ii) were able to provide informed consent, (iii) were fluent in English, (iv) presented with anxious-misery symptoms (anxious-misery group), or (v) had no history of mental health issues or diagnoses (control group).

Participants were excluded if they met any of the following conditions: (i) were currently pregnant; (ii) were unable to tolerate, or else were medically or surgically contraindicative to MR scanning procedures; (iii) presented with any significant handicaps that would interfere with testing procedures; (iv) showed signs of cognitive impairment (MOCA < 24) or neurological disorders; (v) had a history of alcohol use disorder, schizophrenia, or any psychotic disorder(s).

To incorporate a wide range of symptomatology within the anxious-misery group, participants’ eligibility was determined by scoring one standard deviation above the population mean (≥26.2 for males, ≥30.1 for females) on neuroticism on the Neuroticism-Extraversion-Openness Five-Factor Inventory (NEOFFIN). Neuroticism was chosen as the criterion as it correlates highly with the general aspects of psychopathology seen across the spectrum of disorders of interest, namely depression, anxiety, and trauma.

The resulting cohort (N = 241) consisted of participants recruited from within the University of Pennsylvania and the surrounding community. 47 of these participants constituted the control group of individuals who did not meet criteria for any DSM-V diagnosis and scored less than one standard deviation above the population means listed above. Of those 241, 160 identified as female, 73 identified as male, three identified as something other than the options listed, three did not report an answer, and two identified as transgender. Ages ranged from 18 – 59 years old (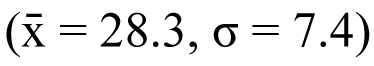. Years of education ranged from 9 – 20 years (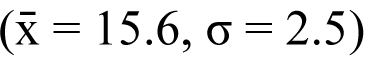.

Participants were not required to cease taking any psychoactive medications. A majority of participants (73.4%) were unmedicated. All participants signed an informed consent form, and the protocol was approved by the Institutional Review Board for human subject research at the University of Pennsylvania. The authors assert that all procedures contributing to this work comply with the ethical standards of the relevant national and institutional committees on human experimentation and with the Helsinki Declaration of 1975, as revised in 2008.

### Measures

Data included 149 clinical items (see Table 1) and 12 neurocognitive test scores measured by the items taken from both the University of Pennsylvania Computerized Neurocognitive Test Battery (Gur et al., 2010) as well as the National Institute of Health Toolbox (Weintraub, 2013).

**Table 1.**
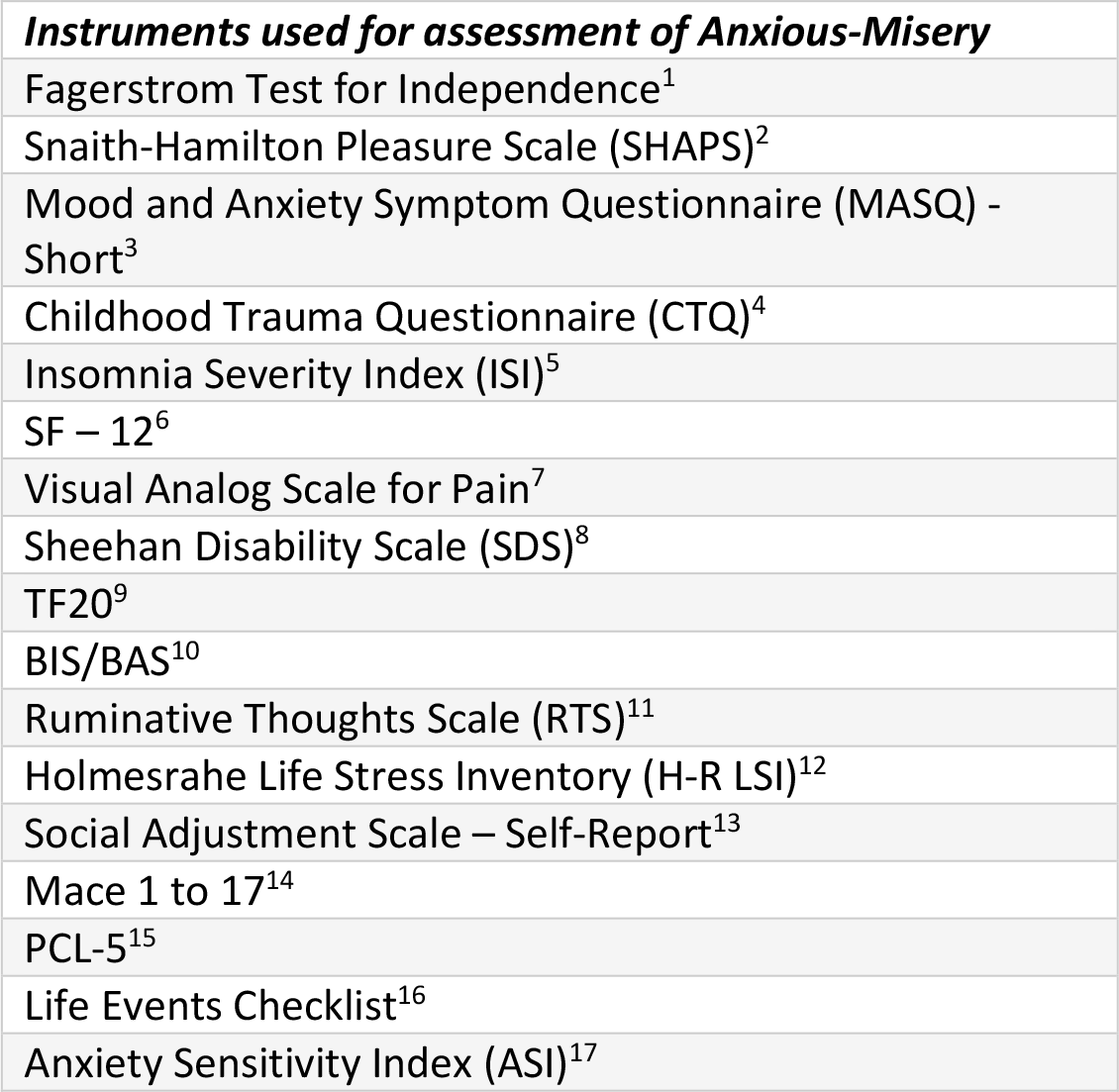
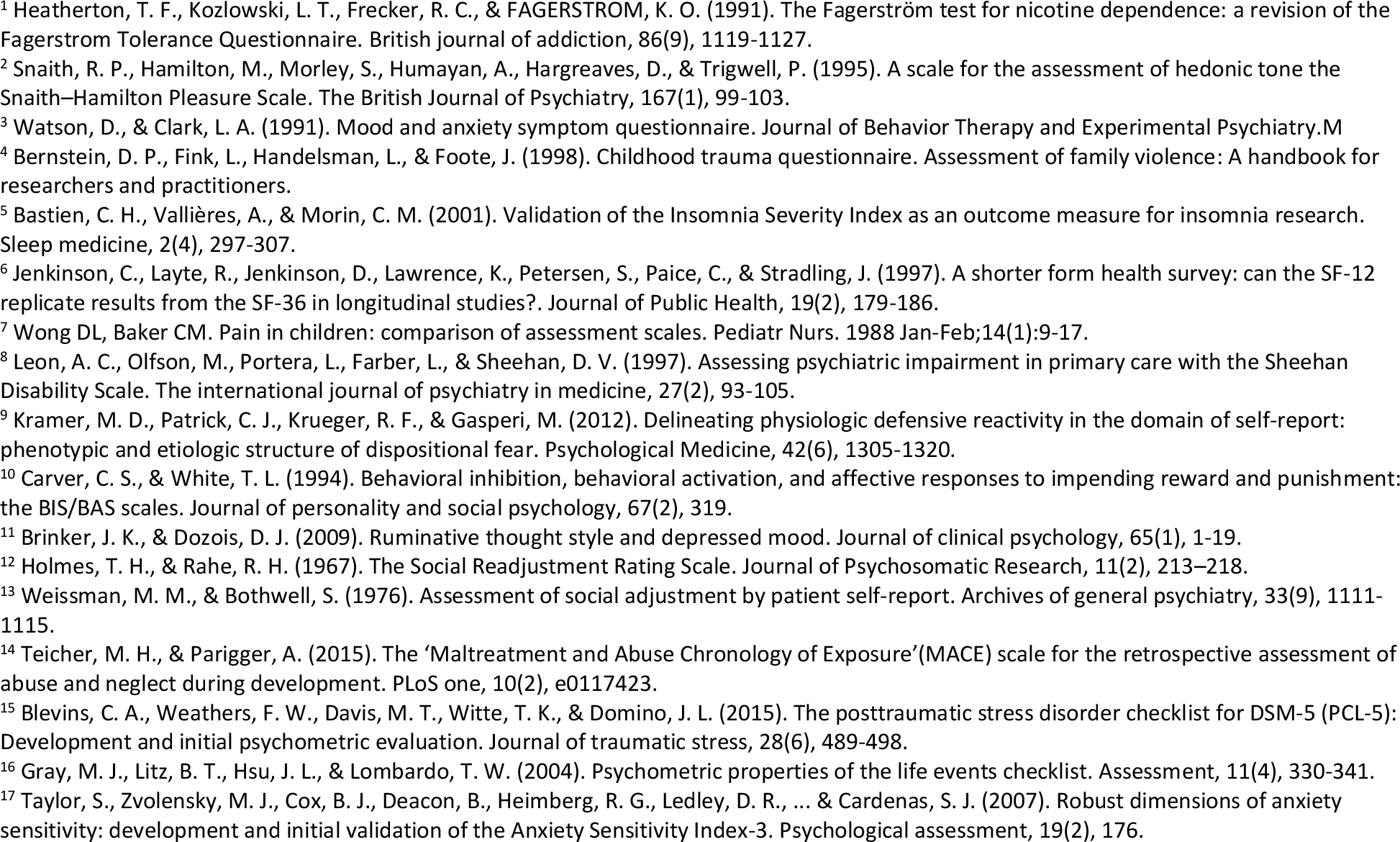
Full list of assessments used in factor analysis.

### Statistical Analysis

The first step was to explore the latent structure of the clinical items using exploratory factor analysis (EFA). Because the items included a mix of Likert-type responses ranging from three (e.g. HAMD general somatic symptoms) to seven (e.g. MADRS inner tension) response options, the inter-item correlation matrix used for analysis included a mix of types, depending on the variable combination. That is, correlations between two continuous items (6 or 7 options) were Pearson correlations; correlations between two ordinal items (<6 options) were polychoric; and correlations between continuous and ordinal items were polyserial.

#### Exploratory model

A model was estimated where the number of factors to extract (5) was determined by a combination of interpretability, subjective evaluation of the scree plot, and fit of the exploratory model. The fit of the model was determined using the Comparative Fit Index (CLI; >0.90 acceptable), Tucker-Lewis Index (TLI; >0.90 acceptable), root mean-square error of approximation (RSMEA; <0.08 acceptable), and standardized root mean-square residual (SRMR; <0.08 acceptable) (Hu & Bentler, 1999). With the correlation matrix estimated and number of factors determined, the factor structure was obtained using least-squares extraction and promax rotation.

#### Confirmatory (CFA) bifactor model

The second step in analysis was to estimate a confirmatory (CFA) bifactor model (Reise, 2012; Reise, Moore, & Haviland, 2010) in which each item loaded on both a general factor (comprising all items) and a specific factor corresponding to the cluster to which it was assigned by the exploratory analysis above. This resulted in a total of six factors (general, plus the five defined above), where a benefit of the bifactor model is that inclusion of the general factor obviates the need for inter-factor correlations, making all latent factors orthogonal.

In the same structural equation model as the CFA, twelve neurocognitive test scores were included as dependent variables relating to the latent clinical factors in the CFA. That is, each neurocognitive test score was predicted by all six factors in the bifactor measurement model, and because the six factors were orthogonal, collinearity was not a concern. Fit of this model was evaluated using the same indices as listed above, and the standardized effects between the clinical and cognitive variables were interpreted. Note that the goal of these analyses was not to test the latent structure of psychopathology, in which case the data would need to be split into separate samples for exploratory and confirmatory analyses. Instead, the goal here was merely to define a model that would fit best to this sample, where the effects of interest were not the measurement model itself, but rather the relationships between the optimally defined clinical dimensions and the neurocognitive test scores.

## Results

Table 2 shows the results of the exploratory factor analysis of the 149 items, with promax rotation. The first factor, which we call “Anxiety”, is most strongly determined by BISBAS items “I worry about making mistakes” and “I feel worried when I think I have done poorly at something important”. The second factor (“Depression”) is most strongly determined by the HAMD suicide item and MADRS suicidal thoughts item. The third factor (“Insomnia”) is most strongly determined by the MADRS reduced sleep item and ISI item “How (dis)satisfied are you with your current sleep pattern?”. The fourth factor (“Impulsivity”) is most strongly determined by the BISBAS items “When I go after something I use a ‘no holds barred’ approach” and “I crave excitement and new sensations”. Finally, the fifth factor (“Somatizing”) is most strongly determined by MASQ item “Had trouble swallowing” and ASI item “When my stomach is upset, I worry that I might be seriously ill”. Inter-factor correlations were mostly moderate (mean = 0.41; range = 0.12-0.67), suggesting the existence of a robust general factor.

Figure 3 shows the results of the structural equation model including the bifactor measurement model described above, as well as twelve neurocognitive tests. Fit of the model is mostly acceptable, with a CFI of 0.93, RMSEA of 0.033±0.002, and SRMR of 0.085. The paths from the measurement model to the cognitive scores are limited to those reaching statistical significance (p < 0.05), though all paths (from all factors to all tests) were included in the model. Note also that items in the measurement portion of Figure xx are truncated (items 3, 6, 9, 12, and 15 are place-holders) for ease of visualization. For a complete list of effects for the measurement model and SEM, see Supplementary Tables 4 and 5. The largest effects to note are from Somatizing to Word Memory, Picture Vocabulary, and List Sorting (Std. β = -0.21, -0.17, & - 0.16, respectively); Impulsivity to Matrix Reasoning (Std. β = 0.16); Insomnia to Picture Sequence (Std. β = -0.16); and Depression to Delay Discounting (Std. β = -0.16). Notably, all effects from Anxiety and Impulsivity were in directions suggesting higher clinical scores were associated with better performance.

To further explore the positive association between symptoms and cognition described above, we split the full sample into their individual diagnostic groups (i.e. MDD, GAD, PTSD, & Control) and tested for mean differences in cognition among those groups. The diagnostic groups were compared using one-way ANOVA, revealing no significant differences on any of the outcome measures. This did not reveal any significant group differences in neurocognitive performance. In an attempt to maximize signal, the same analyses were repeated by diagnostic group, but included only the top ¼ of participants in each group measured by symptom severity on MADRS (MDD), MASQ (GAD), and LEC5 (PTSD). This revealed one significant result in those with MDD symptomatology and their performance on the NIH Toolbox List Sorting task (Weintraub, 2013), which measures working memory performance (p < 0.05). Once again, this result showed that it was the symptomatic participants who scored significantly higher (indicative of better WM performance) compared to the control cohort (see Figure 1).

**Figure 1.**
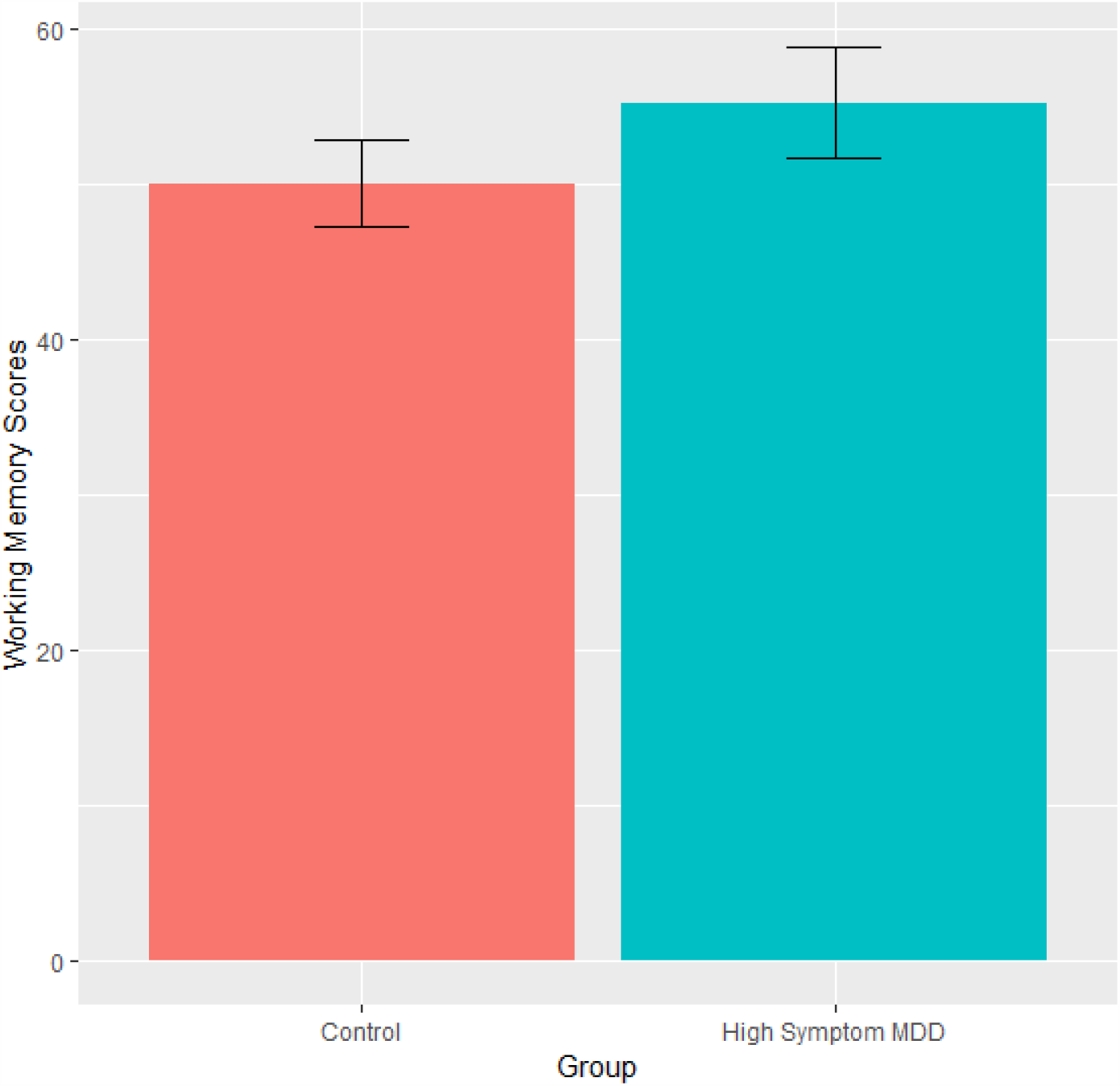
Performance on NIH Toolbox List Sorting Working Memory Task.

## Discussion

These results show that in general, higher levels of clinical symptomatology tended to correlate with better performance across multiple neurocognitive domains. The ‘Impulsivity’ factor was associated with higher performance on assessments measuring general IQ, while the ‘Anxiety’ factor was associated with higher scores on long-term memory, working memory, and executive functioning. ‘Insomnia’ only had one negative association with performance on episodic memory. Depression, as a factor, had one significant association with the Delay Discounting task, with symptomatic patients showing increased likelihood of delaying reward as compared to control.

In contrast to the general trend for higher performance associated with higher levels of psychopathology, ‘Somatizing’ was the only factor associated with worse performance. This included worse performance on memory (both working and episodic), as well as lower performance on general intelligence measures, for symptomatic participants compared with healthy controls.

While these results may at first seem surprising based on the commonly evidenced finding that higher levels of clinical anxiety and depression symptoms are generally associated with worse performance on domains such as working memory and executive function, there is previous literature which supports our results.

In a previous study looking at cognitive performance across multiple domains within a transdiagnostic psychotic population, researchers found a similar effect of higher depression scores associated with better performance. While high levels of psychosis were associated with lower cognitive scores across diagnoses, higher scores on a latent depression factor (while controlling for diagnosis) were associated with better performance on a variety of social cognition tasks (Service et al., 2020). Similar findings were found in previous work that measured depression as a factor within co-morbid psychotic disorders as well; with sub-threshold depression levels showing better performance than controls in terms of accuracy on cognitive tasks. This effect persisted even after controlling for symptom severity and pre-morbid intelligence (Chiappelli et al., 2014).

Another study llooking at a transdiagnostic group of participants, performed a similar analysis to the one used in this paper. Symptomology was grouped (using EFA) into four distinct subfactors, similar to the five we used here. These factors included distress, fear, psychoticism, and externalizing. In addition, they also used the Penn Computerized Neurobehavioral Test Battery to measure performance across similar cognitive domains as the ones described in this study (general intelligence, memory, working memory, etc.). Interestingly, while controlling for “p”, or the general level of psychopathology, they found that the ‘distress’ factor was associated with better performance across all cognitive domains measured (Jonas et al., in press). This is strikingly similar to our results which seem to imply that when diagnosis and symptom severity are controlled for, factors traditionally thought to correlate with lower cognitive performance tend to have less, or even opposite, correlations with cognitive performance.

One theory to potentially explain these unexpected results is that there are complex relationships among intelligence levels, ruminative cognition, and anxious and depressive symptomatology, potentially resulting in this counterintuitive directionality of symptomatology and cognitive performance. Navrady found that those with higher neuroticism were at higher risk for MDD diagnosis as well as self-reported depression, and additionally found that higher levels of intelligence within the same sample were at higher risk of depression once neuroticism levels had been controlled for (Navrady et al., 2017). Another study found that participants with high levels of neuroticism who also had high levels of emotional intelligence were able to successfully mediate their levels of task anxiety better than those with low emotional intelligence (Smith, Saklofske, & Nordstokke, 2014). Verbal intelligence specifically has been found to correlate with higher levels of rumination in a non-depressed sample (Penney, Miedema, & Mazmanian, 2015). This link between general intelligence and ruminative cognition has been proposed before in the Analytical Rumination Hypothesis (Andrews & Thomson, 2009), which suggested that rumination may actually be an adaptive strategy of the mind that is meant to increase the number of cognitive resources allocated to problem solving, which can subsequently result in the cardinal symptoms of depression.

Rumination has long been proposed as a relevant factor mediating how depression might affect some individuals compared to others (Nolen-Hoeksema, 1991). Specifically, people who are shown to have high levels of rumination are found to have both longer episodes of depression as well as more severe episodes. (Nolen-Hoeksema, 1991; 1993). Rumination has been shown to have a variable relationship with depression which may not be as straightforward as previously thought. For example, it has been evidenced that rumination significantly correlates with higher levels of intelligence when the shared variance of rumination with depressive symptomatology is controlled for (du Pont et al., 2020). Rumination has also been found to mediate neuroticism, potentially explaining why some people with high levels of neuroticism develop mental illness while others do not, though the link is still only partially understood (Roberts, Gilboa, & Gotlib, 1998; Hervas & Vazquez, 2011). Due to these correlations, it may be that we were picking up on this proposed correlation of high intelligence with rumination.

However, it must be said that research in this area has also shown contrary results; that high levels of neuroticism are correlated with lower levels of intelligence. One such experiment looked at how the NEOFFIN traits corelated with performance on general intelligence measures and found, as they had predicted, that those with higher neuroticism did worse than their peers with low N, though they do note that this may be due to testing anxiety (Moutafi, Furnham & Paltiel, 2005). Another study even finds evidence that while ‘general’ neuroticism does indeed correlate with lower intelligence levels, certain subfactors of neuroticism (e.g., worry/vulnerability) correlate with higher intelligence (Hill et al., 2020).

A limitation of this study that should be acknowledged includes the use of a sample of convenience, that included many college students from the University of Pennsylvania and surrounding Philadelphia colleges and universities, which may have skewed towards individuals with higher IQs in both the anxious-misery and control conditions, but as such would not necessarily explain the direction of differences between the groups.

In summary, in this cohort we found that participants with higher levels of neuroticism scores significantly outperformed participants with lower neuroticism on cognitive tasks measuring short and long-term memory, as well as executive functioning. Previous literature has found evidence for the possibility of general intelligence being significantly correlated with levels of neuroticism which could lead to increased cognitive performance. This potential connection between the two is further evidenced here, but as noted above, other studies have found opposite results, showing that heightened neuroticism correlates more with lower IQ. Further research is needed to disentangle these contradictory findings, and future experiments could focus on directly examining the link between IQ and neuroticism, and, if necessary, controlling for variable IQ levels in the populations that are studied.

## Data Availability

All data produced in the present study are available upon reasonable request to the authors.

## Author contributions

CRediT author statement according to: https://www.elsevier.com/authors/policies-and-guidelines/credit-author-statement.

**Conceptualization**: **YIS**

**Methodology**: **YIS**,

**TM Software**: **TM**

**Formal analysis**: **FN, TM**

**Investigation**: **FN, TM, JCS, WM, YS**

**Writing - Original Draft**: **FN, TM**

**Writing - Review & Editing**: **FN, YS, TM, JCS**

**Visualization**: **FN, TM**

**Supervision**: **YIS**

**Project administration**: **YIS**

**Funding acquisition**: **YIS**

## Ethical Standards

The authors assert that all procedures contributing to this work comply with the ethical standards of the relevant national and institutional committees on human experimentation and with the Helsinki Declaration of 1975, as revised in 2008.

