## Supplement for "Neuroticism levels in severe depression may reveal association with higher levels of intelligence"

Polychoric and polyserial correlations are approximations of the correlation between the latent variables assumed to be underlying the observed variables. Polychorics are unnecessary for two continuous variables because the observed variables themselves are assumed to be distributed approximately the same as the latent variables, so the correlation between the latent variables underlying them is assumed to be the same as the (Pearson) correlation between the observed variables. However, for ordinal variables, it is not possible for the observed variable (item) to be distributed like a continuous latent variable because there are not enough possible values for the observed distribution to approximate a continuous distribution. The most extreme case is an observed item with only two values (e.g. 0/1 for yes/no), where a binary distribution is clearly not continuous, but the same applies to Likert items with only a few response options. It is assumed that a continuous latent variable underlies the ordinal observed variable, but the distribution of the ordinal variable cannot approximate the latent continuous variable because there are too few response options.

Polychoric correlations approximate the correlation between the latent variables underlying two ordinal variables by using the contingency table between the ordinal variables. The algorithm used for calculation finds what the correlation between the two latent variables would need to be to produce the contingency table in question. For polyserial correlations, a table of means of the continuous variable by values of the ordinal variable is used rather than a contingency table, but the logic is the same: the algorithm finds what the correlation between the continuous variable and the latent trait underlying the ordinal variable would need to be to produce that table of means.

Table 2. Exploratory Factor Analysis of 149 Clinical Items.

| Item | Anxiety | Depression | Insomnia | Impulsivity | Somatizing |
|---|---|---|---|---|---|
| bisbas_makemistakes | 0.94 | | | | |
| bisbas_poorly | 0.93 | | | | |
| bisbas_angryatme | 0.87 | | | | |
| rts_whenihave | 0.79 | | | | |
| rts_ifihave | 0.74 | | | | |
| rts_wheni3 | 0.71 | | | | |
| rts_ifind3 | 0.71 | | | | |
| bisbas_nofear | 0.71 | | | | |
| rts_wheni | 0.70 | | | | |
| bisbas_workedup | 0.69 | | | | |
| rts_wheni2 | 0.69 | | | | |
| rts_ifind | 0.68 | | | | |
| rts_ifthere | 0.64 | | | | |
| rts_itend | 0.64 | | | | |
| bisbas_scolding | 0.62 | | | | |
| neo_worrier_v2 | -0.61 | | | | |
| asi_sweat | 0.59 | | | | |
| asi_tremble | 0.57 | | | | |
| bisbas_fewfears | 0.57 | | | 0.41 | |
| rts_sometimese | 0.57 | | | | |
| rts_whentrying | 0.56 | | | | |
| neo_rarely_fearful_v2 | -0.56 | | | | |
| rts_icant | 0.56 | | | | |
| rts_ifind2 | 0.54 | | | | |
| asi_notice | 0.53 | | | | 0.41 |
| masq_worry | 0.52 | | | | |
| rts_wheniaml | 0.51 | | | | |
| rts_wheniam | 0.50 | | | | |
| rts_ihave | 0.49 | | | | |
| masq_felt_nervous | 0.48 | | | | |
| neo_going_to_pieces_v2 | 0.47 | 0.45 | | | |
| masq_trembling | 0.45 | | | | |
| neo_want_to_hide_v2 | 0.45 | 0.39 | | | |
| rts_sometimes | 0.44 | | | | |
| asi_blush | 0.42 | | | | |
| asi_ontaskscared | 0.40 | | | | 0.38 |
| rts_evenif | 0.40 | 0.31 | | | |
| rts_itisvery | 0.36 | | | | |

| | | | | | |
|---|---|---|---|---|---|
| asi_appearnervous | 0.34 | | | | 0.34 |
| masq_shakyhands | 0.33 | | | | |
| neo_tense_jittery_v2 | 0.32 | | | | |
| masq_racingheart | 0.28 | | | | |
| rts_ilike | 0.27 | | | | |
| bisbas_haircut | 0.23 | | | | |
| hamd_suicide | | 0.98 | | | |
| madrs_suicidal_thoughts | | 0.88 | | | |
| madrs_pessimistic_thoughts | | 0.87 | | | |
| hamd_feelings_of_guilt | | 0.87 | | | |
| masq_felt_hopeless | | 0.85 | | | |
| madrs_apparent_sadness | | 0.74 | | | |
| masq_felt_discouraged | | 0.73 | | | |
| hamd_depressed_mood | | 0.73 | | | |
| hamd_retardation | -0.30 | 0.73 | | | |
| hamd_insight | -0.38 | 0.70 | -0.35 | | 0.55 |
| masq_felt_worthless | | 0.70 | | | |
| masq_failure | | 0.67 | | | |
| madrs_reported_sadness | | 0.67 | | | |
| masq_disappointed | | 0.65 | | | |
| masq_felt_sad | | 0.64 | | | |
| madrs_inability_to_feel | | 0.63 | | | |
| neo_worthless_v2 | 0.36 | 0.63 | | | |
| masq_blamed_self | | 0.59 | | | |
| hamd_loss_of_weight_by_history | | 0.57 | | | |
| masq_feltgood | | 0.54 | | 0.32 | |
| masq_lookforward | | 0.53 | | | |
| neo_discouraged_v2 | 0.37 | 0.51 | | | |
| hamd_work_and_activities | | 0.51 | 0.37 | | |
| madrs_concentration_difficulties | | 0.50 | | | |
| neo_helpless_v2 | | 0.48 | | | |
| masq_proud | | 0.48 | | | |
| masq_confident | 0.37 | 0.47 | | | |
| hamd_somatic_symptoms_gi | | 0.47 | | | |
| neo_lonely_blue_v2 | -0.40 | -0.47 | | | |
| masq_accomplished_shitton | | 0.44 | | | |
| neo_angry_people_treat_me_v2 | | 0.43 | | | |
| madrs_lassitude | | 0.43 | 0.36 | | |
| neo_seldom_sad_v2 | -0.32 | -0.41 | | | |
| masq_interestingthings | | 0.41 | | 0.31 | |
| neo_bitter_v2 | | 0.39 | | | |

| | | | | |
|---|---|---|---|---|
| madrs_inner_tension | 0.38 | | | |
| madrs_reduced_appetite | 0.35 | | | |
| hamd_agitation | 0.26 | | | |
| bisbas_familyisimportant | 0.24 | | | |
| madrs_reduced_sleep | | 0.79 | | |
| isi_isi_4 | | 0.78 | | |
| hamd_insomnia_early | | 0.77 | | |
| hamd_insomnia_late | -0.32 | 0.77 | | |
| isi_isi_6 | | 0.76 | | |
| isi_early | | 0.75 | | |
| isi_isi_5 | | 0.74 | | |
| isi_middle | | 0.73 | | |
| isi_isi_7 | | 0.72 | | |
| hamd_insomnia_middle | | 0.66 | | |
| isi_late | | 0.58 | | |
| shaps_enjoyprogram | | 0.53 | | |
| shaps_enjoydrink | | 0.52 | | |
| shaps_enjoyreading | | 0.48 | | |
| shaps_enjoylookingsmart | | 0.47 | | |
| shaps_enjoybath | | 0.46 | 0.31 | |
| hamd_genital_symptoms | | 0.44 | | |
| shaps_enjoysmells | | 0.41 | | |
| hamd_somatic_symptoms_general | 0.35 | 0.40 | | |
| shaps_enjoypraise | | 0.40 | | |
| shaps_enjoyhelpingothers | | 0.39 | | |
| shaps_pleasureinhobbies | 0.31 | 0.39 | 0.36 | |
| shaps_enjoysmallthings | | 0.38 | 0.37 | |
| shaps_enjoymeal | | 0.38 | 0.37 | |
| hamd_anxiety_somatic | | 0.37 | | 0.32 |
| shaps_enjoyfamily | 0.34 | 0.35 | | |
| shaps_enjoyview | | 0.34 | 0.31 | |
| hamd_hypochondriasis | | 0.23 | | |
| bisbas_noholds | | | 0.76 | |
| bisbas_crave | | | 0.75 | |
| bisbas_opportunity | | | 0.73 | |
| bisbas_trynew | | | 0.72 | |
| bisbas_wantsomething | | | 0.66 | |
| bisbas_want | | | 0.66 | |
| bisbas_goodthings | | | 0.65 | |
| bisbas_allout | | | 0.65 | |
| bisbas_mightbefun | | | 0.65 | |

| | | | | | |
|---|---|---|---|---|---|
| bisbas_moveonit | | | | 0.62 | |
| bisbas_spur | | -0.47 | | 0.59 | |
| masq_felt_happy | | | | 0.42 | |
| bisbas_wincontest | | | | 0.40 | |
| bisbas_doingwell | | 0.31 | | 0.39 | |
| shaps_enjoysmilingfaces | | 0.30 | | 0.39 | |
| masq_energy | | | | 0.34 | |
| bisbas_peopleact | | | | -0.32 | |
| masq_lively | | | | 0.31 | |
| bisbas_dress | | | | 0.28 | |
| masq_swallowing | | | | | 0.68 |
| asi_stomach | | | | | 0.65 |
| asi_tightchest | | | | | 0.64 |
| asi_throat | | -0.38 | | | 0.64 |
| asi_skipbeat | | | | | 0.62 |
| asi_heart | 0.32 | | | | 0.59 |
| asi_thoughtsspeed | 0.38 | | | | 0.54 |
| asi_heartattack | | | | | 0.54 |
| masq_twitch | | | 0.44 | | 0.51 |
| asi_mindblank | 0.30 | | | | 0.50 |
| asi_spacey | | | | | 0.50 |
| asi_troublethinking | 0.37 | | | | 0.44 |
| masq_shortofbreath | | | | | 0.43 |
| asi_faintpublic | | | | | 0.43 |
| masq_felt_numb_in_body | | | 0.32 | | 0.40 |
| asi_ontaskcrazy | 0.34 | | | | 0.38 |
| masq_drymouth2 | | | | | 0.35 |
| masq_dizzy | | | | | 0.33 |
| hamd_anxiety_psychic | | | | | 0.31 |

Inter-Factor Correlations

| | F1 | F2 | F3 | F4 | F5 |
|---|---|---|---|---|---|
| F1 | 1.00 | 0.56 | 0.46 | 0.18 | 0.52 |
| F2 | 0.56 | 1.00 | 0.67 | 0.42 | 0.42 |
| F3 | 0.46 | 0.67 | 1.00 | 0.39 | 0.36 |
| F4 | 0.18 | 0.42 | 0.39 | 1.00 | 0.12 |
| F5 | 0.52 | 0.42 | 0.36 | 0.12 | 1.00 |

Note: Loadings with absolute value < 0.30 removed unless the primary loading on a factor.

Table 3. Confirmatory Bifactor Analysis of 149 Clinical Items.

| Item | General | Anxiety | Depression | Insomnia | Impulsivity | Somatizing |
|---|---|---|---|---|---|---|
| bisbas_makemistakes | 0.46 | 0.62 | 0 | 0 | 0 | 0 |
| bisbas_poorly | 0.45 | 0.61 | 0 | 0 | 0 | 0 |
| bisbas_angryatme | 0.39 | 0.57 | 0 | 0 | 0 | 0 |
| rts_whenihave | 0.51 | 0.65 | 0 | 0 | 0 | 0 |
| rts_ifihave | 0.45 | 0.65 | 0 | 0 | 0 | 0 |
| rts_wheni3 | 0.54 | 0.55 | 0 | 0 | 0 | 0 |
| rts_ifind3 | 0.58 | 0.56 | 0 | 0 | 0 | 0 |
| bisbas_nofear | 0.37 | 0.47 | 0 | 0 | 0 | 0 |
| rts_wheni | 0.69 | 0.65 | 0 | 0 | 0 | 0 |
| bisbas_workedup | 0.55 | 0.47 | 0 | 0 | 0 | 0 |
| rts_wheni2 | 0.56 | 0.58 | 0 | 0 | 0 | 0 |
| rts_ifind | 0.67 | 0.66 | 0 | 0 | 0 | 0 |
| rts_ifthere | 0.62 | 0.52 | 0 | 0 | 0 | 0 |
| rts_itend | 0.49 | 0.58 | 0 | 0 | 0 | 0 |
| bisbas_scolding | 0.40 | 0.39 | 0 | 0 | 0 | 0 |
| neo_worrier_v2 | -0.45 | -0.40 | 0 | 0 | 0 | 0 |
| asi_sweat | 0.50 | 0.41 | 0 | 0 | 0 | 0 |
| asi_tremble | 0.39 | 0.46 | 0 | 0 | 0 | 0 |
| bisbas_fewfears | 0.35 | 0.36 | 0 | 0 | 0 | 0 |
| rts_sometimese | 0.53 | 0.46 | 0 | 0 | 0 | 0 |
| rts_whentrying | 0.65 | 0.38 | 0 | 0 | 0 | 0 |
| neo_rarely_fearful_v2 | -0.63 | -0.33 | 0 | 0 | 0 | 0 |
| rts_icant | 0.72 | 0.61 | 0 | 0 | 0 | 0 |
| rts_ifind2 | 0.71 | 0.61 | 0 | 0 | 0 | 0 |
| asi_notice | 0.58 | 0.41 | 0 | 0 | 0 | 0 |
| masq_worry | 0.78 | 0.28 | 0 | 0 | 0 | 0 |
| rts_wheniaml | 0.28 | 0.52 | 0 | 0 | 0 | 0 |
| rts_wheniam | 0.64 | 0.40 | 0 | 0 | 0 | 0 |
| rts_ihave | 0.56 | 0.36 | 0 | 0 | 0 | 0 |
| masq_felt_nervous | 0.65 | 0.32 | 0 | 0 | 0 | 0 |
| neo_going_to_pieces_v2 | 0.71 | 0.17 | 0 | 0 | 0 | 0 |
| masq_trembling | 0.49 | 0.32 | 0 | 0 | 0 | 0 |
| neo_want_to_hide_v2 | 0.75 | 0.18 | 0 | 0 | 0 | 0 |
| rts_sometimes | 0.63 | 0.35 | 0 | 0 | 0 | 0 |
| asi_blush | 0.35 | 0.35 | 0 | 0 | 0 | 0 |
| asi_ontaskscared | 0.70 | 0.22 | 0 | 0 | 0 | 0 |
| rts_evenif | 0.74 | 0.34 | 0 | 0 | 0 | 0 |
| rts_itisvery | 0.70 | 0.31 | 0 | 0 | 0 | 0 |

| | | | | | | |
|---|---|---|---|---|---|---|
| asi_appearnervous | 0.37 | 0.30 | 0 | 0 | 0 | 0 |
| masq_shakyhands | 0.41 | 0.24 | 0 | 0 | 0 | 0 |
| neo_tense_jittery_v2 | 0.68 | 0.22 | 0 | 0 | 0 | 0 |
| masq_racingheart | 0.56 | 0.21 | 0 | 0 | 0 | 0 |
| rts_ilike | -0.03 | 0.34 | 0 | 0 | 0 | 0 |
| bisbas_haircut | 0.42 | 0.12 | 0 | 0 | 0 | 0 |
| hamd_suicide | 0.35 | 0 | 0.84 | 0 | 0 | 0 |
| madrs_suicidal_thoughts | 0.45 | 0 | 0.72 | 0 | 0 | 0 |
| madrs_pessimistic_thoughts | 0.72 | 0 | 0.41 | 0 | 0 | 0 |
| hamd_feelings_of_guilt | 0.67 | 0 | 0.38 | 0 | 0 | 0 |
| masq_felt_hopeless | 0.79 | 0 | 0.42 | 0 | 0 | 0 |
| madrs_apparent_sadness | 0.72 | 0 | 0.41 | 0 | 0 | 0 |
| masq_felt_discouraged | 0.82 | 0 | 0.31 | 0 | 0 | 0 |
| hamd_depressed_mood | 0.73 | 0 | 0.43 | 0 | 0 | 0 |
| hamd_retardation | 0.41 | 0 | 0.41 | 0 | 0 | 0 |
| hamd_insight | 0.29 | 0 | 0.29 | 0 | 0 | 0 |
| masq_felt_worthless | 0.79 | 0 | 0.39 | 0 | 0 | 0 |
| masq_failure | 0.78 | 0 | 0.31 | 0 | 0 | 0 |
| madrs_reported_sadness | 0.76 | 0 | 0.38 | 0 | 0 | 0 |
| masq_disappointed | 0.78 | 0 | 0.27 | 0 | 0 | 0 |
| masq_felt_sad | 0.81 | 0 | 0.26 | 0 | 0 | 0 |
| madrs_inability_to_feel | 0.68 | 0 | 0.34 | 0 | 0 | 0 |
| neo_worthless_v2 | 0.76 | 0 | 0.18 | 0 | 0 | 0 |
| masq_blamed_self | 0.80 | 0 | 0.18 | 0 | 0 | 0 |
| hamd_loss_of_weight_by_history | 0.29 | 0 | 0.46 | 0 | 0 | 0 |
| masq_feltgood | 0.83 | 0 | 0.30 | 0 | 0 | 0 |
| masq_lookforward | 0.69 | 0 | 0.42 | 0 | 0 | 0 |
| neo_discouraged_v2 | 0.75 | 0 | 0.10 | 0 | 0 | 0 |
| hamd_work_and_activities | 0.74 | 0 | 0.26 | 0 | 0 | 0 |
| madrs_concentration_difficulties | 0.70 | 0 | 0.20 | 0 | 0 | 0 |
| neo_helpless_v2 | 0.71 | 0 | 0.10 | 0 | 0 | 0 |
| masq_proud | 0.75 | 0 | 0.30 | 0 | 0 | 0 |
| masq_confident | 0.81 | 0 | 0.25 | 0 | 0 | 0 |
| hamd_somatic_symptoms_gi | 0.35 | 0 | 0.70 | 0 | 0 | 0 |
| neo_lonely_blue_v2 | -0.70 | 0 | -0.05 | 0 | 0 | 0 |
| masq_accomplished_shitton | 0.65 | 0 | 0.28 | 0 | 0 | 0 |
| neo_angry_people_treat_me_v2 | 0.54 | 0 | 0.06 | 0 | 0 | 0 |
| madrs_lassitude | 0.72 | 0 | 0.23 | 0 | 0 | 0 |
| neo_seldom_sad_v2 | -0.67 | 0 | -0.03 | 0 | 0 | 0 |
| masq_interestingthings | 0.61 | 0 | 0.44 | 0 | 0 | 0 |
| neo_bitter_v2 | 0.58 | 0 | 0.02 | 0 | 0 | 0 |

| | | | | | | |
|---|---|---|---|---|---|---|
| madrs_inner_tension | 0.80 | 0 | -0.06 | 0 | 0 | 0 |
| madrs_reduced_appetite | 0.42 | 0 | 0.63 | 0 | 0 | 0 |
| hamd_agitation | 0.33 | 0 | 0.06 | 0 | 0 | 0 |
| bisbas_familyisimportant | 0.21 | 0 | 0.01 | 0 | 0 | 0 |
| madrs_reduced_sleep | 0.48 | 0 | 0 | 0.58 | 0 | 0 |
| isi_isi_4 | 0.60 | 0 | 0 | 0.60 | 0 | 0 |
| hamd_insomnia_early | 0.41 | 0 | 0 | 0.56 | 0 | 0 |
| hamd_insomnia_late | 0.40 | 0 | 0 | 0.45 | 0 | 0 |
| isi_isi_6 | 0.55 | 0 | 0 | 0.65 | 0 | 0 |
| isi_early | 0.51 | 0 | 0 | 0.54 | 0 | 0 |
| isi_isi_5 | 0.52 | 0 | 0 | 0.59 | 0 | 0 |
| isi_middle | 0.48 | 0 | 0 | 0.47 | 0 | 0 |
| isi_isi_7 | 0.62 | 0 | 0 | 0.65 | 0 | 0 |
| hamd_insomnia_middle | 0.44 | 0 | 0 | 0.43 | 0 | 0 |
| isi_late | 0.47 | 0 | 0 | 0.35 | 0 | 0 |
| shaps_enjoyprogram | 0.50 | 0 | 0 | 0.53 | 0 | 0 |
| shaps_enjoydrink | 0.60 | 0 | 0 | 0.57 | 0 | 0 |
| shaps_enjoyreading | 0.62 | 0 | 0 | 0.47 | 0 | 0 |
| shaps_enjoylookingsmart | 0.34 | 0 | 0 | 0.52 | 0 | 0 |
| shaps_enjoybath | 0.52 | 0 | 0 | 0.51 | 0 | 0 |
| hamd_genital_symptoms | 0.51 | 0 | 0 | 0.28 | 0 | 0 |
| shaps_enjoysmells | 0.61 | 0 | 0 | 0.42 | 0 | 0 |
| hamd_somatic_symptoms_general | 0.63 | 0 | 0 | 0.22 | 0 | 0 |
| shaps_enjoypraise | 0.42 | 0 | 0 | 0.45 | 0 | 0 |
| shaps_enjoyhelpingothers | 0.57 | 0 | 0 | 0.33 | 0 | 0 |
| shaps_pleasureinhobbies | 0.69 | 0 | 0 | 0.39 | 0 | 0 |
| shaps_enjoysmallthings | 0.67 | 0 | 0 | 0.42 | 0 | 0 |
| shaps_enjoymeal | 0.61 | 0 | 0 | 0.42 | 0 | 0 |
| hamd_anxiety_somatic | 0.55 | 0 | 0 | 0.09 | 0 | 0 |
| shaps_enjoyfamily | 0.58 | 0 | 0 | 0.37 | 0 | 0 |
| shaps_enjoyview | 0.72 | 0 | 0 | 0.41 | 0 | 0 |
| hamd_hypochondriasis | 0.36 | 0 | 0 | 0.05 | 0 | 0 |
| bisbas_noholds | 0.17 | 0 | 0 | 0 | 0.72 | 0 |
| bisbas_crave | 0.15 | 0 | 0 | 0 | 0.68 | 0 |
| bisbas_opportunity | 0.31 | 0 | 0 | 0 | 0.73 | 0 |
| bisbas_trynew | 0.46 | 0 | 0 | 0 | 0.60 | 0 |
| bisbas_wantsomething | 0.28 | 0 | 0 | 0 | 0.68 | 0 |
| bisbas_want | 0.22 | 0 | 0 | 0 | 0.71 | 0 |
| bisbas_goodthings | 0.14 | 0 | 0 | 0 | 0.61 | 0 |
| bisbas_allout | 0.26 | 0 | 0 | 0 | 0.74 | 0 |
| bisbas_mightbefun | 0.22 | 0 | 0 | 0 | 0.62 | 0 |

| | | | | | | |
|---|---|---|---|---|---|---|
| bisbas_moveonit | 0.36 | 0 | 0 | 0 | 0.68 | 0 |
| bisbas_spur | -0.02 | 0 | 0 | 0 | 0.52 | 0 |
| masq_felt_happy | 0.62 | 0 | 0 | 0 | 0.30 | 0 |
| bisbas_wincontest | 0.25 | 0 | 0 | 0 | 0.41 | 0 |
| bisbas_doingwell | 0.35 | 0 | 0 | 0 | 0.40 | 0 |
| shaps_enjoysmilingfaces | 0.69 | 0 | 0 | 0 | 0.26 | 0 |
| masq_energy | 0.73 | 0 | 0 | 0 | 0.28 | 0 |
| bisbas_peopleact | 0.42 | 0 | 0 | 0 | -0.32 | 0 |
| masq_lively | 0.47 | 0 | 0 | 0 | 0.26 | 0 |
| bisbas_dress | 0.19 | 0 | 0 | 0 | 0.30 | 0 |
| masq_swallowing | 0.29 | 0 | 0 | 0 | 0 | 0.42 |
| asi_stomach | 0.46 | 0 | 0 | 0 | 0 | 0.58 |
| asi_tightchest | 0.40 | 0 | 0 | 0 | 0 | 0.70 |
| asi_throat | 0.36 | 0 | 0 | 0 | 0 | 0.68 |
| asi_skipbeat | 0.41 | 0 | 0 | 0 | 0 | 0.68 |
| asi_heart | 0.55 | 0 | 0 | 0 | 0 | 0.51 |
| asi_thoughtsspeed | 0.63 | 0 | 0 | 0 | 0 | 0.50 |
| asi_heartattack | 0.38 | 0 | 0 | 0 | 0 | 0.66 |
| masq_twitch | 0.44 | 0 | 0 | 0 | 0 | 0.29 |
| asi_mindblank | 0.63 | 0 | 0 | 0 | 0 | 0.47 |
| asi_spacey | 0.63 | 0 | 0 | 0 | 0 | 0.49 |
| asi_troublethinking | 0.72 | 0 | 0 | 0 | 0 | 0.47 |
| masq_shortofbreath | 0.45 | 0 | 0 | 0 | 0 | 0.35 |
| asi_faintpublic | 0.46 | 0 | 0 | 0 | 0 | 0.35 |
| masq_felt_numb_in_body | 0.53 | 0 | 0 | 0 | 0 | 0.25 |
| asi_ontaskcrazy | 0.70 | 0 | 0 | 0 | 0 | 0.34 |
| masq_drymouth2 | 0.38 | 0 | 0 | 0 | 0 | 0.20 |
| masq_dizzy | 0.50 | 0 | 0 | 0 | 0 | 0.26 |
| hamd_anxiety_psychic | 0.72 | 0 | 0 | 0 | 0 | 0.07 |

Table 4. Standardized Effect Sizes from Structural Equation Model Relating Psychopathology to Cognitive Tests.

| DV | IV | Std. Effect | p-value |
|---|---|---|---|
| Card Sort | **PUNISHMENT** | **0.124** | **0.049** |
| | ANXIOUSMIS | -0.082 | 0.179 |
| | INSOMNIA | -0.013 | 0.815 |
| | IMPULSIVE | 0.010 | 0.886 |
| | SOMATIZING | -0.058 | 0.404 |
| | GENERAL | -0.029 | 0.659 |
| Flanker | PUNISHMENT | 0.071 | 0.275 |
| | ANXIOUSMIS | 0.007 | 0.911 |
| | INSOMNIA | -0.011 | 0.856 |
| | IMPULSIVE | 0.004 | 0.958 |
| | SOMATIZING | -0.022 | 0.758 |
| | GENERAL | -0.034 | 0.598 |
| List Sorting | PUNISHMENT | -0.042 | 0.498 |
| | ANXIOUSMIS | 0.000 | 0.996 |
| | INSOMNIA | 0.095 | 0.124 |
| | IMPULSIVE | 0.118 | 0.062 |
| | **SOMATIZING** | **-0.163** | **0.033** |
| | GENERAL | 0.061 | 0.321 |
| Pattern Comparison | **PUNISHMENT** | **0.142** | **0.018** |
| | ANXIOUSMIS | -0.019 | 0.756 |
| | INSOMNIA | -0.030 | 0.607 |
| | IMPULSIVE | 0.062 | 0.310 |
| | SOMATIZING | -0.025 | 0.726 |
| | GENERAL | -0.014 | 0.822 |
| Picture Sequence | **PUNISHMENT** | **0.110** | **0.046** |
| | ANXIOUSMIS | -0.074 | 0.206 |
| | **INSOMNIA** | **-0.161** | **0.002** |
| | IMPULSIVE | 0.060 | 0.336 |
| | SOMATIZING | -0.070 | 0.345 |
| | GENERAL | 0.057 | 0.364 |
| Picture Vocabulary | PUNISHMENT | 0.067 | 0.282 |
| | ANXIOUSMIS | -0.058 | 0.326 |
| | INSOMNIA | -0.001 | 0.984 |
| | **IMPULSIVE** | **0.152** | **0.008** |
| | **SOMATIZING** | **-0.167** | **0.033** |
| | **GENERAL** | **0.133** | **0.026** |

| | | | |
|---|---|---|---|
| Delay Discounting | PUNISHMENT | 0.035 | 0.582 |
| | **ANXIOUSMIS** | **-0.157** | **0.004** |
| | INSOMNIA | 0.012 | 0.833 |
| | IMPULSIVE | 0.055 | 0.369 |
| | SOMATIZING | 0.082 | 0.237 |
| | GENERAL | -0.045 | 0.466 |
| Word Memory | PUNISHMENT | -0.008 | 0.887 |
| | ANXIOUSMIS | -0.093 | 0.109 |
| | INSOMNIA | -0.018 | 0.778 |
| | IMPULSIVE | -0.022 | 0.721 |
| | **SOMATIZING** | **-0.205** | **0.005** |
| | GENERAL | 0.080 | 0.199 |
| Matrix Reasoning | PUNISHMENT | -0.023 | 0.709 |
| | ANXIOUSMIS | -0.024 | 0.693 |
| | INSOMNIA | 0.001 | 0.982 |
| | **IMPULSIVE** | **0.163** | **0.012** |
| | SOMATIZING | -0.124 | 0.099 |
| | GENERAL | 0.035 | 0.564 |
| Emotion Identification | PUNISHMENT | 0.012 | 0.836 |
| | ANXIOUSMIS | -0.093 | 0.103 |
| | INSOMNIA | -0.017 | 0.786 |
| | IMPULSIVE | 0.066 | 0.297 |
| | SOMATIZING | -0.052 | 0.512 |
| | GENERAL | 0.037 | 0.531 |
| Trails A Time | **PUNISHMENT** | **-0.118** | **0.018** |
| | ANXIOUSMIS | 0.048 | 0.520 |
| | INSOMNIA | 0.037 | 0.600 |
| | IMPULSIVE | 0.046 | 0.499 |
| | SOMATIZING | 0.082 | 0.249 |
| | GENERAL | 0.028 | 0.653 |
| Trails B Efficiency | PUNISHMENT | -0.057 | 0.383 |
| | ANXIOUSMIS | 0.019 | 0.801 |
| | INSOMNIA | -0.009 | 0.892 |
| | IMPULSIVE | 0.067 | 0.323 |
| | SOMATIZING | -0.103 | 0.104 |
| | GENERAL | -0.036 | 0.556 |

Note. Significant effects bolded; DV = dependent variable; IV = independent variable.

Figure 5. Structural Equation Model Combining the Bifactor Measurement Model of Clinical Items with the Individual Cognitive Tests as Dependent Variables.

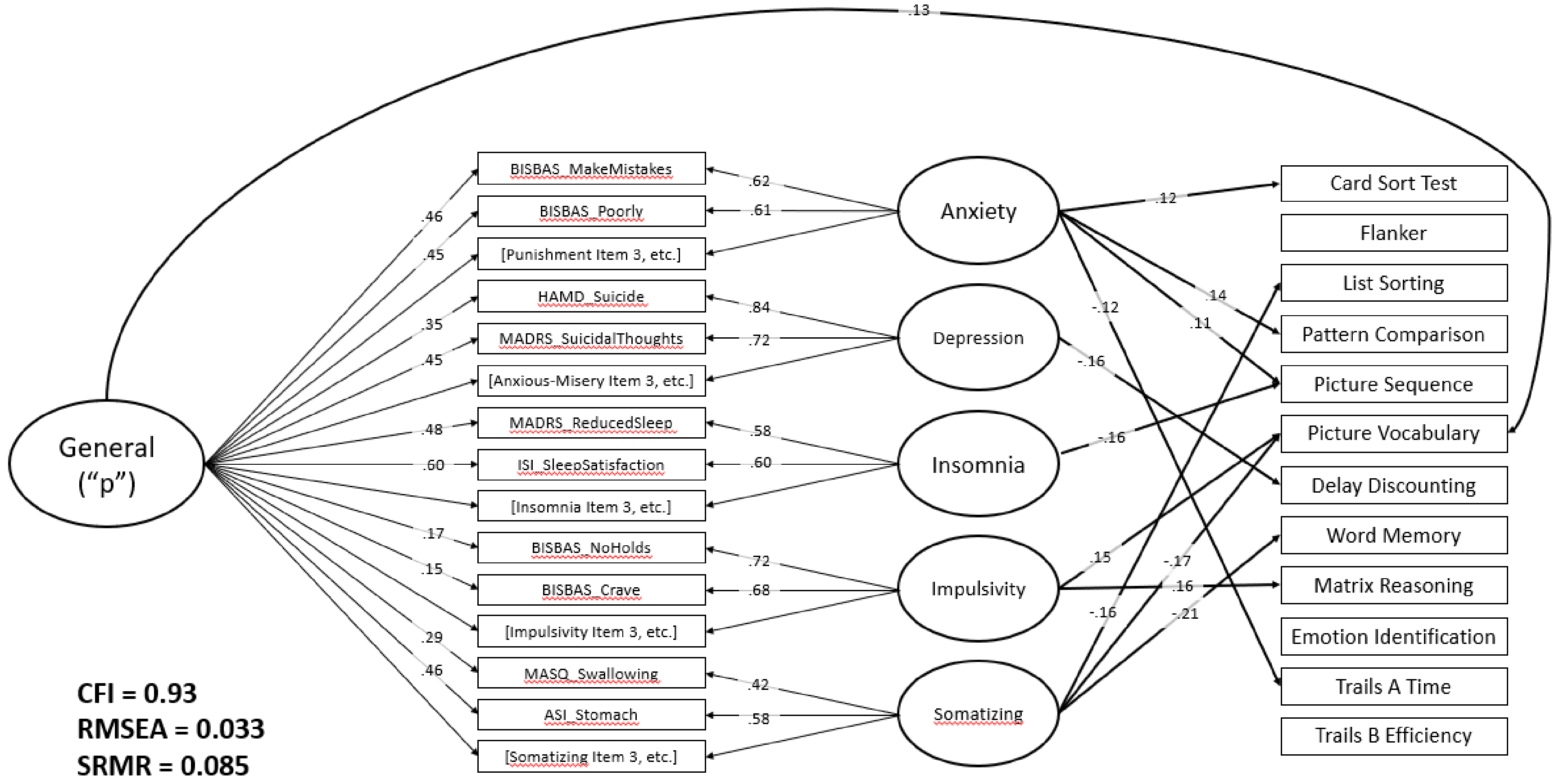
